# Effects of polygenic liability for autism on neonatal thalamocortical connectivity and behavioral outcomes across sex

**DOI:** 10.64898/2025.12.23.25342934

**Authors:** Emily Chiem, Sai Sruthi Amirtha Ganesh, Jack Dodson, Mirella Dapretto, Leanna Hernandez

**Affiliations:** Molecular, Cellular, and Integrative Physiology Program, University of California, Los Angeles, Los Angeles, CA 90095, USA; Ahmanson-Lovelace Brain Mapping Center, University of California, Los Angeles, Los Angeles, CA 90095, USA; Center for Autism Research and Treatment, University of California, Los Angeles, Los Angeles, CA 90095, USA; Department of Psychiatry and Biobehavioral Sciences, University of California, Los Angeles, Los Angeles, CA 90095, USA

## Abstract

Functional brain networks are altered in Autism Spectrum Disorder (ASD), with differences in thalamocortical connectivity detectable as early as infancy. ASD shows distinct sex differences, not only in diagnostic rates, but also in brain and behavioral manifestations of the condition. Although common variants account for much of the genetic liability for ASD, little is known about the impact of ASD-associated genetic variation on functional brain connectivity and behavioral outcomes in early life or how this may differ between males and females. Here, we utilize functional MRI (fMRI), genetic, and behavioral data from the Developing Human Connectome Project (dHCP) to investigate sex differences in the association between ASD polygenic scores (PGS), thalamocortical functional connectivity (37-44 weeks postmenstrual age), and behavioral outcomes (18 months) in European term-born infants. We show that across the full sample, higher ASD PGS is associated with weaker thalamic connectivity with posterior parietal cortex, as well as greater ASD-related and ADHD symptoms and slower motor development. Sex differences in the relationship between ASD PGS and thalamic connectivity largely encompassed sensorimotor, posterior parietal, temporal, and insular cortices. Further, in female infants, thalamic connectivity patterns associated with greater genetic liability for ASD were related to poorer motor development. These findings suggest genetic predisposition for ASD shapes early thalamocortical functional connectivity in a sex-specific manner and negatively impacts behavioral development in early toddlerhood.

## Introduction

Autism Spectrum Disorder (ASD) is rarely diagnosed before the age of two [1], with an average age of diagnosis of five years [2]. Importantly, however, alterations in functional brain connectivity are present as early as infancy, indicating that the neurodevelopmental origins of ASD begin well before clinical diagnosis (e.g., [3], [4]). Infants with a family history of ASD are at about 10-times greater risk of developing the disorder relative to the general population [5] and show detectable alterations in thalamocortical connectivity at 6 postnatal weeks, which are associated with social attention and sensory challenges [6], [7]. Primary sensory networks, such as connectivity between the thalamus and cortex, are among the earliest emerging brain networks, setting the stage for the development of higher-order socioemotional and cognitive networks [8]. The thalamus plays a key role in the gating of sensory inputs to the cortex [9] and in the modulation of corticocortical signaling [10] – circuits which have known alterations in ASD [11], [12], [13]. Emerging evidence has also implicated the thalamus as an early site of genetic susceptibility in ASD. For example, a recent transcriptomic study reported that the developing fetal thalamus expresses high levels of ASD-associated genes [14]. Taken together, these findings suggest that thalamic circuitry may be an important nexus of vulnerability in ASD.

Common inherited genetic variation accounts for an estimated 50% of liability for ASD [15]. Indeed, recent genome wide association studies (GWAS) have detected 12 variants associated with the disorder, indicating a highly polygenic genetic architecture [16]. Leveraging these GWAS findings, polygenic scores (PGS) can be calculated to estimate an individual’s cumulative genetic liability for ASD relative to others in the population. Prior neuroimaging studies have largely focused on the relationship between ASD PGS and structural brain measures. For example, typically developing children ages 3-14 with higher ASD PGS show greater cortical thickness in the precentral and postcentral gyri, as well as the precuneus [17]. Emerging work indicates that, even in the neonatal period, higher ASD PGS is associated with alterations in brain structure, including increased fronto-temporal and reduced parieto-occipital brain volumes [18], as well as greater white matter fiber-bundle cross section [19]. Furthermore, higher ASD PGS has been associated with behavioral outcomes, such as poorer motor and language development in infants [20], [21], and behavioral regulation difficulties in children and adolescents [22]. A major goal of pediatric neuroscience is to identify early brain-based biomarkers of neurodevelopmental disorders, yet how ASD-associated genetic variation influences neonatal functional brain connectivity and subsequent behavioral trajectories remain to be elucidated.

In addition to genetic and neurodevelopmental factors, sex plays a significant role in ASD risk. ASD is diagnosed three to four times more often in males than females [23], a striking difference hypothesized to be driven in part by the female protective effect, which posits that females require a greater genetic load to display ASD symptoms [24]. Overall, sex differences in functional brain connectivity have been reported across several brain networks [25], [26], [27], including thalamic connectivity [28] in autistic individuals. Prior work also suggests that the downstream effects of ASD genetic liability on brain connectivity may differ by sex; ASD-associated variants are associated with altered salience network connectivity in male, but not female adolescents with and without ASD [29] and show opposite sex-specific patterns of effect on reward network connectivity [30]. However, it remains unclear how the genome-wide polygenic liability for ASD may differentially impact male and female brain development in early infancy.

To address this gap, we took a multimodal approach, leveraging large-scale functional MRI (fMRI), genetic, and behavioral data from the Developing Human Connectome Project (dHCP) to investigate how genetic predisposition for ASD influences neonatal thalamic functional connectivity. ASD PGS were derived using the largest published GWAS of ASD and tested for associations with thalamocortical connectivity at 37-44 weeks postmenstrual age, as well as later behavioral outcomes at 18 months. Given the known sex-bias in ASD diagnoses, we also test for potential sex differences in these effects. By integrating imaging, genetics, and behavior in infancy, this work aims to define early neural pathways impacted by polygenic liability for ASD to help inform the development of effective strategies for early detection and intervention.

## Methods

### Participants

Data were obtained from neonatal participants recruited from a population-based sample at the St Thomas’ Hospital, London as part of the Developing Human Connectome Project (dHCP; www.developingconnectome.org). The original dHCP data collection included a total of 783 infants with MRI data, 800 infants with genetic data, and 627 infants with behavioral data. Written parental consent was obtained for each infant. Inclusion criteria for the dHCP study included infants between 23-44 weeks gestational age estimated from mother’s last menstrual period and confirmed where possible with an ultrasound scan. Exclusion criteria included infants with contraindication to MR imaging (e.g., implanted metallic devices) or inadequate communication during the consent process to parents due to language barriers. The dHCP study was approved by the United Kingdom Health Research Authority [31]. For the current study, we included term-born (gestational age ≥ 37 weeks) infants scanned between 37-44 weeks postmenstrual age, determined to have European ancestry, as described below, yielding a final sample size of N_PGS+MRI_ = 271; N_PGS+behavior_ = 348-349.

### Genotyping

Infant saliva samples were collected using the Oragene DNA OG-250 kits (DNAGenoteck Inc., Kanata, Canada) at both the initial neonatal MRI visit and the 18-month follow up visit. One sample (usually the first) per infant was genotyped by the NIHR BioResource Centre Maudsley Genomics & Biomarker Core Facility on the Illumina Infinium Omni5-4 array v1.2, resulting in a total of 4,327,108 genotyped single-nucleotide polymorphisms (SNPs). Genotyping was performed in two batches. Basic quality control was performed by the Department of Biostatistics & Health Informatics, King’s College London. Additional processing was performed according to the GenomeStudio quality control Standard Operating Procedure [32], as well as a pipeline which identified and removed samples with call rates below 95% [32], identified gender mismatches, and flagged potential heterozygosity outliers. Additional details regarding genotype quality control and processing can be found in [31].

### MRI data acquisition

Infants were scanned between 37-44 postmenstrual weeks of age at the Evelina Newborn Imaging Centre, Centre for the Developing Brain, King’s College London, United Kingdom. A 3T Philips Achieva scanner and dedicated neonatal 32 channel phased array head coil were used to collect anatomical (T1-weighted and T2-weighted), resting-state functional MRI (fMRI), and diffusion MRI scans during natural sleep, except for four infants who were sedated with chloral hydrate. The current study used 15-minute resting-state fMRI scans collected using multiband 9X accelerated echo-planar imaging (TE = 38 ms, TR = 392 ms, 2300 volumes, 2.15 mm isotropic spatial resolution).

### Behavioral follow-up

A battery of parent-reported questionnaires and standardized child-centered assessments were collected at the 18-month follow-up visit. Here, we focused on measures indexing behavioral, communication, and motor-related traits, which are predicted to have an association with ASD PGS: the Quantitative Checklist for Autism in Toddlers (Q-CHAT), a 25-item parent-report questionnaire which is designed for early detection of autistic traits at 18-24 months of age [33]; the Bayley Scales of Infant and Toddler Development, Third Edition (Bayley-III), an individually administered assessment of toddlers’ cognitive, language (receptive and expression), and motor (gross and fine) skills [34]; and the Child Behavior Checklist for Ages 1.5-5 (CBCL), a 100-item parent-report questionnaire designed to assess behavioral and emotional problems [35]. We focused our analyses on the CBCL DSM-Oriented scales (depressive problems, anxiety problems, autism spectrum problems, attention deficit/hyperactivity (ADHD) problems, and oppositional defiant problems) due to previously reported co-occurrences of psychiatric conditions with ASD [36].

### Data Preprocessing and Analysis

#### Genetic Data

SNPs were filtered for minor allele frequency <0.01, missingness per individual >0.01, missingness per marker >0.01, and Hardy-Weinberg equilibrium (p < 10-6) using PLINK v2.0 [37]. Genotype imputation was performed on the Michigan Imputation Server [38] using the TOPMed reference panel [39]. Genetic ancestry was estimated by merging dHCP genotype data with the 1000 Genomes Project European reference panel and applying a K nearest neighbors clustering algorithm. Closely related individuals were excluded from the final sample using a genetic relationship matrix computed using GCTA v1.94 and a filter cutoff of 0.05 [40]. As the genome-wide association study (GWAS) used to generate polygenic scores (PGS) exclusively included European individuals [16], we include only infants of European ancestry in the current analyses.

#### Polygenic scores

Summary statistics were obtained from the largest published GWAS of ASD [16], which included 18,381 individuals with ASD and 27,969 controls of European ancestry. PGS were computed using genotype data from dHCP infants and GWAS summary statistics in PRS-CS, using the European linkage disequilibrium reference panel and a global shrinkage parameter of 1e-2 [41], as recommended for highly polygenic traits.

#### fMRI data

Minimally preprocessed fMRI data was downloaded from the National Institute of Mental Health Data Archive (Collection #3955). Details on the bespoke dHCP fMRI preprocessing pipeline were previously described in [42]. Briefly, preprocessing steps included fieldmap pre-processing using FSL TOPUP [43], motion and susceptibility distortion correction using the FSL EDDY tool [44], high-pass filtering (150s high-pass cutoff), FIX denoising (FMRIB’s ICA-based Xnoiseifier [45]) to remove noise components observed in the spatial independent component analysis (e.g., head movement, multiband artifact, sagittal sinus, arteries, CSF pulsation, and unclassified noise), and nuisance regression of FIX noise independent components and rigid-body motion parameters. The filtered and denoised fMRI data were registered to each subject’s native T2-weigthed space and a standard 40-week template from the dHCP volumetric atlas [46]. Scans were excluded based on dHCP pipeline derived quality control metrics [42], such as mean denoised DVARS outliers (root mean squared intensity difference between successive frames with threshold of 75th percentile + 1.5 times the inter-quartile range), mean denoised temporal signal-to-noise ratio, and registration alignment. Scans with radiology scores of 4 or 5 were also excluded due to incidental findings. We performed additional spatial smoothing (3-mm Gaussian kernel) to improve the signal-to-noise ratio [47], bandpass filtering (0.01 Hz < t < 0.1 Hz) to reduce physiological noise, as well as white matter, CSF, and global signal regression using FSL FEAT.

The bilateral thalamus, derived from the dHCP 40-week template, was used in seed-based resting state functional connectivity analyses. Blood oxygen level dependent (BOLD) time series, averaged across the thalamus, were extracted from processed residuals in standard space and correlated with time series of every other voxel in the brain to generate single-subject functional connectivity maps. Using Fisher’s r-to-z transformation, the resulting correlation maps were converted to z-statistic maps. Group-level analyses were performed in FSL using FMRIB’s Local Analysis of Mixed Effects (FLAME 1+2), with ASD PGS as a bottom-up regressor. In the full sample, the first five ancestry principal components, postmenstrual scan age (weeks), and sex were included as nuisance regressors. Separate analyses were also run in each sex separately using the same nuisance regressors. All contrasts were thresholded at Z > 2.3, and cluster corrected for multiple comparisons at P < 0.05. (Table S1).

#### Behavioral regressions

Statistical analyses were performed in R v4.2.1. The relationship between ASD PGS Z-scores and behavioral measures were tested using the lme4 package. For normally distributed variables (e.g., Q-CHAT total score, Bayley-III composite scores), linear models were fitted, while for non-normally distributed variables (e.g., CBCL t-scores), generalized linear models with gamma distribution and log link were fitted. Covariates included the first 5 ancestry principal components, sex, gestational age at birth, and behavioral assessment age. In addition, we examined the relationship between parameter estimates of sex-specific functional connectivity and behavioral measures found to be significantly associated with ASD PGS. Linear models were fitted for each sex separately with the following covariates: the first 5 ancestry principal components, gestational age at birth, and behavioral assessment age.

## Results

### Polygenic liability for ASD and behavioral profiles in the neonatal dHCP cohort

Infants were recruited from the general population and thus presumed to be neurotypical. There were no significant differences in ASD PGS (t = 0.11, P = 0.91) between sexes. Males had significantly lower scores on the Bayley-III cognitive (t = 2.76, P < 0.01) and language composite scores (t = 3.61, P < 0.001) than females, indicating attenuated motor and language development, but did not differ on any other behavioral measures (Table 1).

**Table 1.**
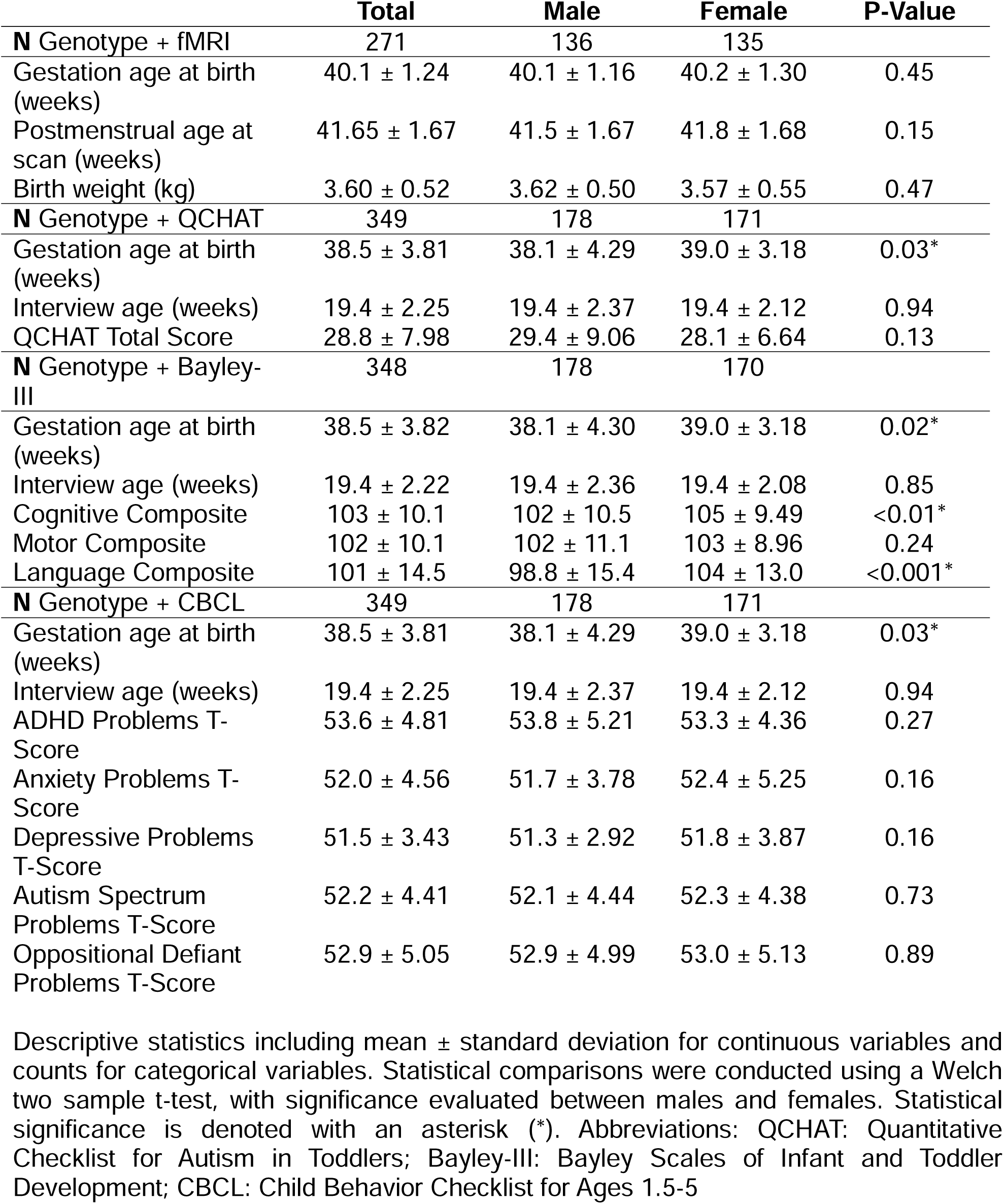
Sample demographics.

|  | **Total** | **Male** | **Female** | **P-Value** |
| --- | --- | --- | --- | --- |
| **N Genotype + fMRI** | **271** | **136** | **135** |  |
| Gestation age at birth (weeks) | 40.1 ± 1.24 | 40.1 ± 1.16 | 40.2 ± 1.30 | 0.45 |
| Postmenstrual age at scan (weeks) | 41.65 ± 1.67 | 41.5 ± 1.67 | 41.8 ± 1.68 | 0.15 |
| Birth weight (kg) | 3.60 ± 0.52 | 3.62 ± 0.50 | 3.57 ± 0.55 | 0.47 |
| **N Genotype + QCHAT** | **349** | **178** | **171** |  |
| Gestation age at birth (weeks) | 38.5 ± 3.81 | 38.1 ± 4.29 | 39.0 ± 3.18 | 0.03* |
| Interview age (weeks) | 19.4 ± 2.25 | 19.4 ± 2.37 | 19.4 ± 2.12 | 0.94 |
| QCHAT Total Score | 28.8 ± 7.98 | 29.4 ± 9.06 | 28.1 ± 6.64 | 0.13 |
| **N Genotype + Bayley-III** | **348** | **178** | **170** |  |
| Gestation age at birth (weeks) | 38.5 ± 3.82 | 38.1 ± 4.30 | 39.0 ± 3.18 | 0.02* |
| Interview age (weeks) | 19.4 ± 2.22 | 19.4 ± 2.36 | 19.4 ± 2.08 | 0.85 |
| Cognitive Composite | 103 ± 10.1 | 102 ± 10.5 | 105 ± 9.49 | <0.01* |
| Motor Composite | 102 ± 10.1 | 102 ± 11.1 | 103 ± 8.96 | 0.24 |
| Language Composite | 101 ± 14.5 | 98.8 ± 15.4 | 104 ± 13.0 | <0.001* |
| **N Genotype + CBCL** | **349** | **178** | **171** |  |
| Gestation age at birth (weeks) | 38.5 ± 3.81 | 38.1 ± 4.29 | 39.0 ± 3.18 | 0.03* |
| Interview age (weeks) | 19.4 ± 2.25 | 19.4 ± 2.37 | 19.4 ± 2.12 | 0.94 |
| ADHD Problems T-Score | 53.6 ± 4.81 | 53.8 ± 5.21 | 53.3 ± 4.36 | 0.27 |
| Anxiety Problems T-Score | 52.0 ± 4.56 | 51.7 ± 3.78 | 52.4 ± 5.25 | 0.16 |
| Depressive Problems T-Score | 51.5 ± 3.43 | 51.3 ± 2.92 | 51.8 ± 3.87 | 0.16 |
| Autism Spectrum Problems T-Score | 52.2 ± 4.41 | 52.1 ± 4.44 | 52.3 ± 4.38 | 0.73 |
| Oppositional Defiant Problems T-Score | 52.9 ± 5.05 | 52.9 ± 4.99 | 53.0 ± 5.13 | 0.89 |

Descriptive statistics including mean ± standard deviation for continuous variables and counts for categorical variables. Statistical comparisons were conducted using a Welch two sample t-test, with significance evaluated between males and females. Statistical significance is denoted with an asterisk (\*). Abbreviations: QCHAT: Quantitative Checklist for Autism in Toddlers; Bayley-III: Bayley Scales of Infant and Toddler Development; CBCL: Child Behavior Checklist for Ages 1.5-5

### ASD PGS are associated with poorer 18-month behavioral outcomes

Models testing the relationship between ASD PGS and later behavioral outcomes identified several significant associations with parent-reported questionnaires and clinician administered assessments at 18 months of age (Figure 1). In the combined male (N = 178) and female (N = 171) sample, higher ASD PGS (reflective of greater genetic predisposition for ASD) was associated with higher scores on the Q-CHAT total scale (β = 0.91, P = 0.04), indicating that elevated genome-wide liability for ASD predicts greater autism traits at 18 months. Higher ASD PGS was also associated with lower scores on the Bayley-III motor composite scale (β = -1.27, P = 0.02, male N = 178, female N = 170), reflecting less developed gross and fine motor skills, and with higher scores on the CBCL ADHD problems scale (β = 0.01, P = 0.03, male N = 178, female N = 171), indicating elevated ADHD-related behavioral problems. All significant gene-behavior relationships showed the same direction of effect in both male and female infants; importantly, however, they did not survive correction for multiple testing (Table S2).

**Figure 1.**
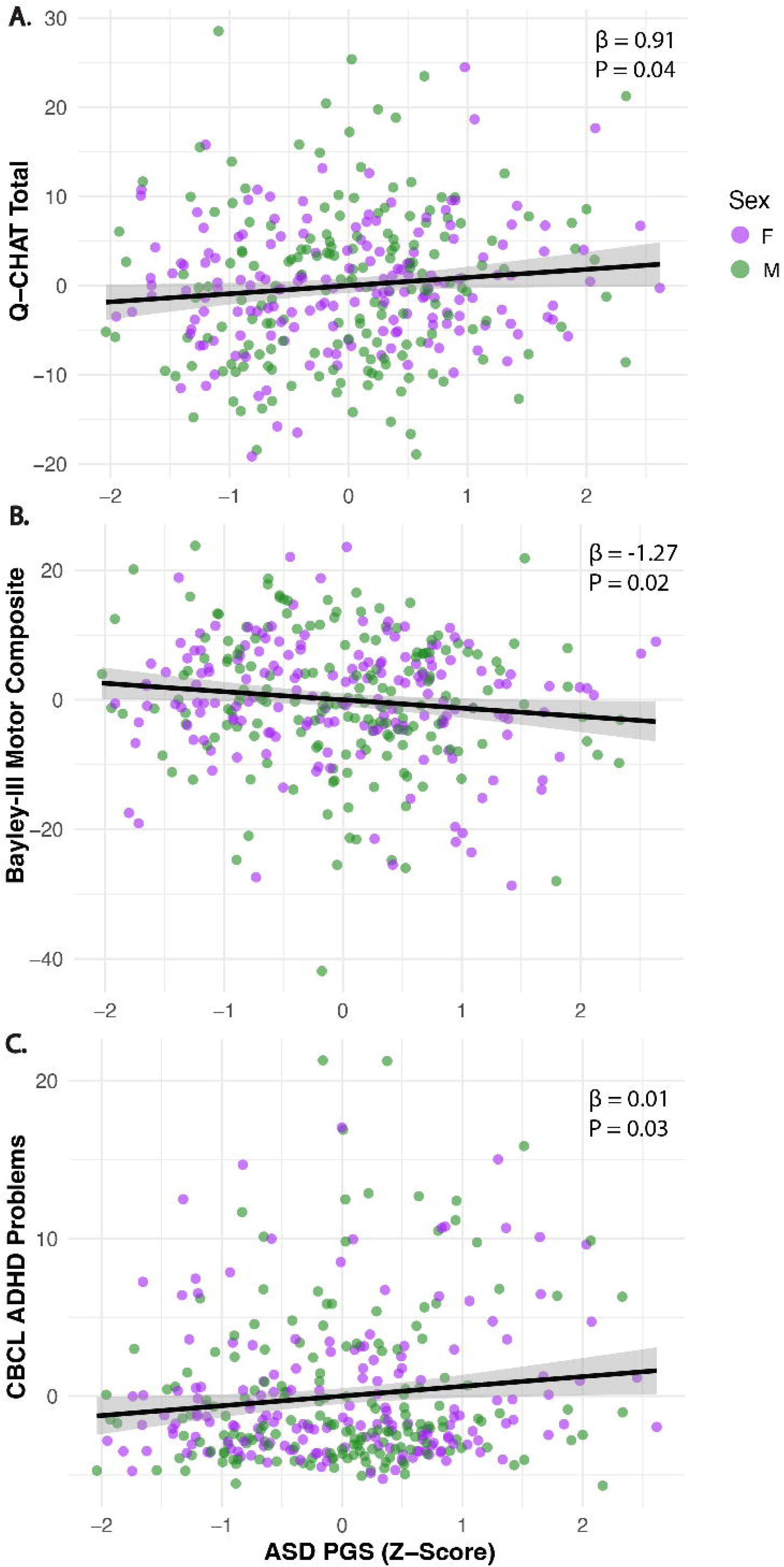
Polygenic scores (PGS) for ASD are associated with behavioral outcomes at 18 months. Higher ASD PGS, which reflects greater genetic predisposition for ASD, significantly predicted A) greater ASD-related symptoms as assessed by the Quantitative Checklist for Autism in Toddlers (Q-CHAT), B) lower motor composite scores on the Bayley Scale of Infant and Toddler Development (Bayley-III), and C) greater ADHD problems on the Child Behavior Checklist (CBCL). Plots show ASD PGS and behavioral measures residualized for covariates (ancestry principal components, sex, assessment age, birth age). Confidence intervals (95%) are indicated by the gray shaded area.

### ASD PGS show sex-specific effects on neonatal thalamocortical connectivity

Next, we examined ASD PGS associations with thalamic functional connectivity at 37- 44 weeks postmenstrual age in the full sample and interactions with sex. In the combined male (N = 136) and female (N = 135) sample, higher ASD PGS was associated with weaker connectivity between the thalamus and right posterior parietal cortex (peak Z = 3.6; Figure 2A, Table S1). In addition, we evaluated sex x ASD PGS interactions in predicting thalamic functional connectivity. Here we found that, relative to female infants, males showed a positive association between ASD PGS and thalamic connectivity with parietal cortex, such that greater genetic liability for ASD predicted stronger connectivity; in contrast, females showed a negative association between ASD PGS and thalamic connectivity with parietal cortex (peak Z = 3.96; Figure 2B, Table S1). A similar pattern emerged in the superior temporal gyrus and insula, where females showed a positive association between ASD PGS and thalamic connectivity, with higher ASD PGS predicting stronger connectivity, whereas males showed a negative association between ASD PGS and thalamic connectivity with these same brain regions (peak Z = 3.43; Figure 2C, Table S1).

**Figure 2.**
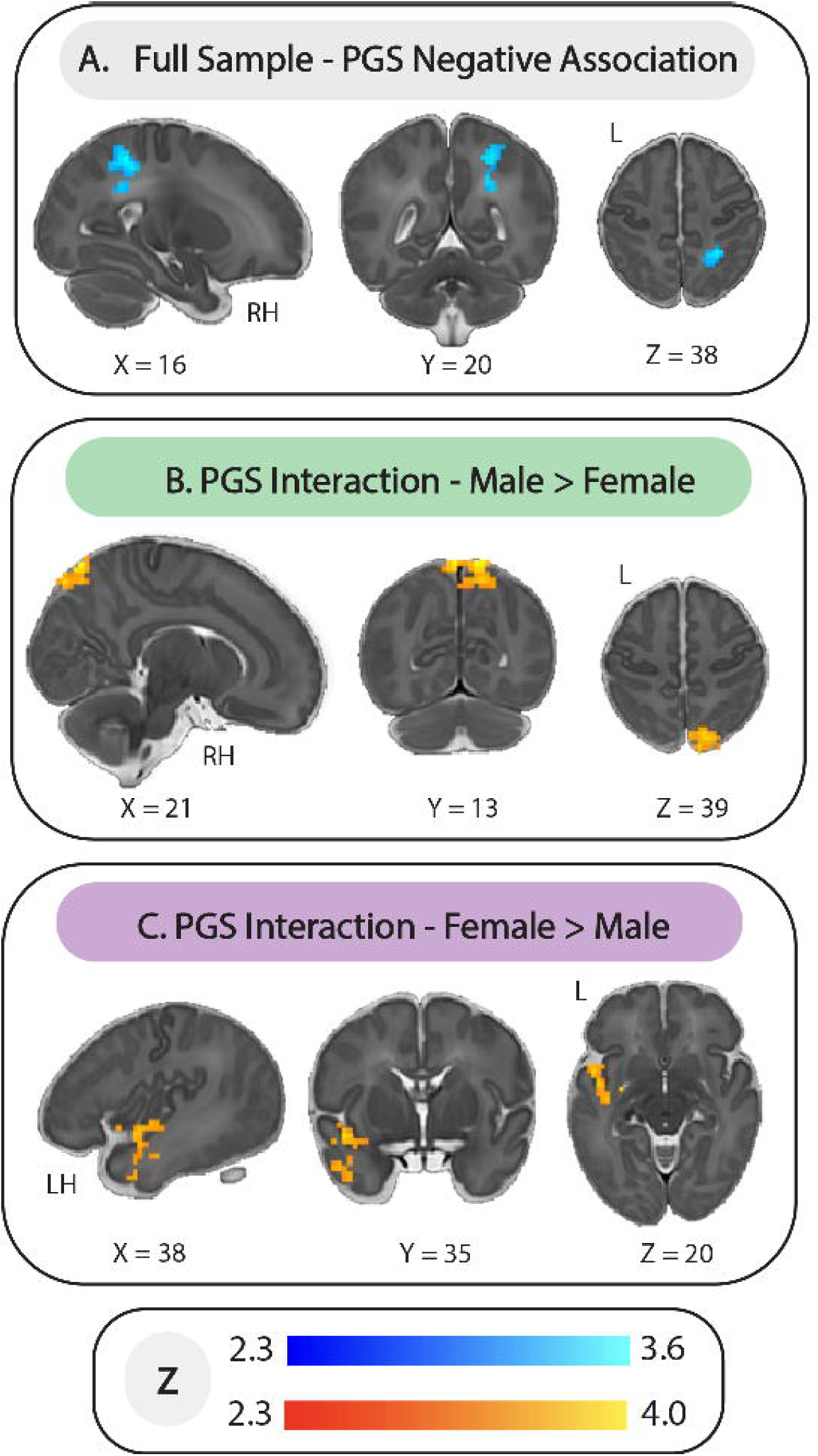
ASD PGS shows sex-differential effects on neonatal thalamocortical connectivity. A) Across the full dHCP sample, higher ASD PGS was associated with weaker thalamic functional connectivity with right posterior parietal cortex in neonates 37-44 weeks postmenstrual age. B) Male infants with higher ASD PGS showed stronger connectivity between thalamus and parietal cortex, while female infants showed weaker thalamic connectivity with the same brain region. C) Female infants with higher ASD PGS showed stronger thalamic connectivity with superior temporal gyrus and insula, while male infants showed weaker thalamic connectivity with these regions as a function of increasing ASD PGS.

The significant sex x ASD PGS interactions next prompted us to examine how the relationship between ASD PGS and thalamic connectivity differed when the sample was stratified by sex. In males, higher ASD PGS was associated with stronger connectivity between the thalamus and left sensorimotor cortex (peak Z = 3.4; Figure 3A, Table S1). Whereas in females, higher ASD PGS was associated with stronger thalamic connectivity with the parieto-cingulate (peak Z = 3.69) and anterior temporal cortices (peak Z = 4.14), as well as weaker thalamic connectivity with the parieto-occipital cortex (peak Z = 3.55; Figure 3B-C, Table S1). Finally, we examined whether these sex-specific patterns of thalamocortical connectivity were associated with behavioral outcomes found to be significantly predicted by ASD PGS (i.e. Q-CHAT total score, Bayley-III motor composite, CBCL ADHD problems). In females only, stronger thalamic connectivity with the anterior temporal cortex was associated with lower scores on the Bayley-III motor composite scale (β = -15.8, P = 0.01; Figure 3B), while stronger thalamic connectivity with the posterior parietal cortex was associated with higher scores on the Bayley-III motor composite scale (β = 12.4, P < 0.01; Figure 3C).

**Figure 3.**
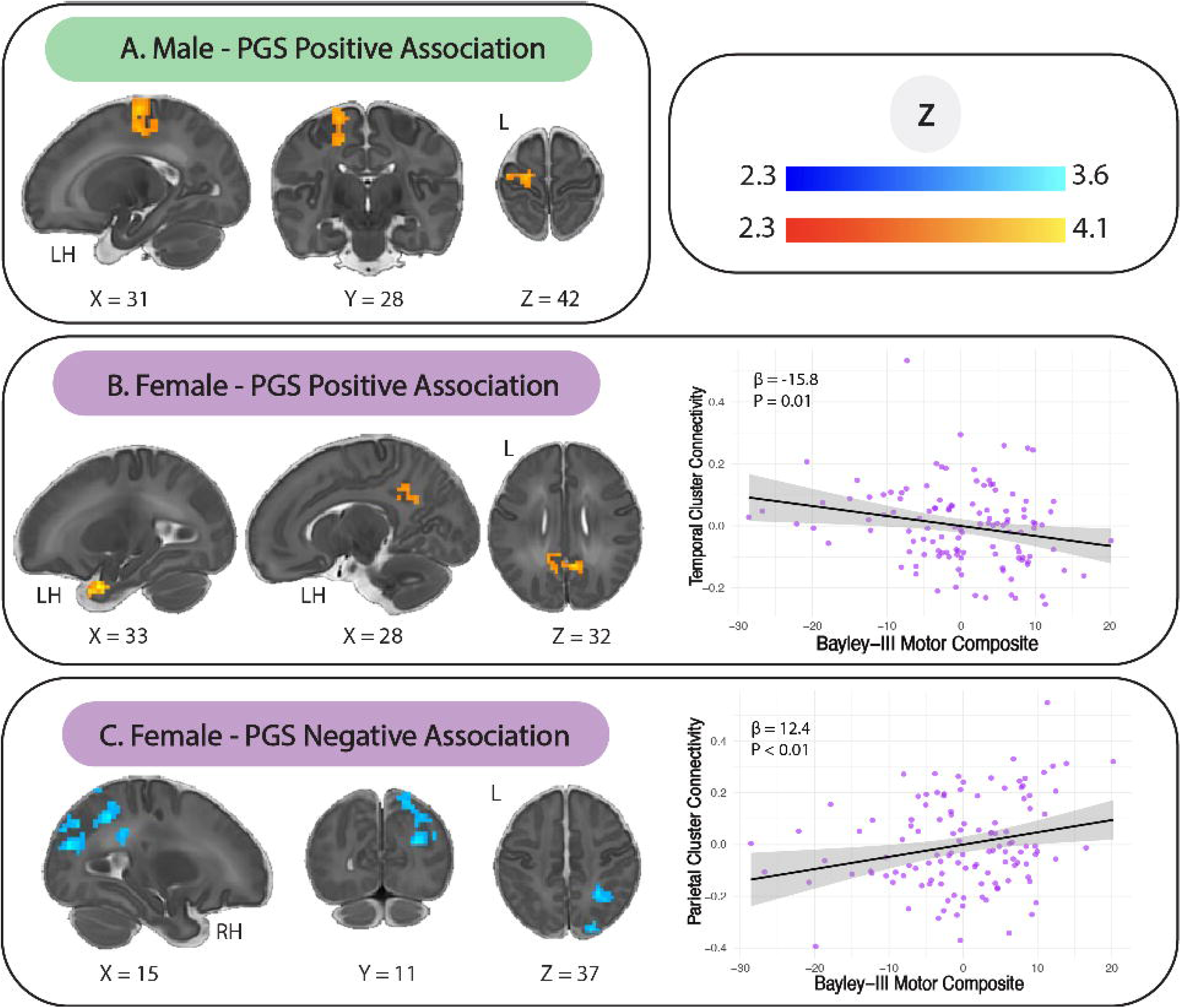
Sex-specific associations between ASD PGS, neonatal thalamic connectivity, and toddler motor development. A) In males, higher ASD PGS was associated with stronger thalamic connectivity with left sensorimotor cortex. B) In females, higher ASD PGS was associated with stronger thalamic connectivity with parieto-cingulate and anterior temporal cortices. Stronger thalamic connectivity with the anterior temporal cortex cluster was related to lower motor development scores at 18 months. C) In females, higher ASD PGS was associated with weaker thalamic connectivity with parieto-occipital cortex. Stronger thalamic connectivity with the parietal cortex cluster was related to higher motor development scores at 18 months. Plots show parameter estimates of functional connectivity and the Bayley-III motor composite residualized for covariates (ancestry principal components, assessment age, birth age). Confidence intervals (95%) are indicated by the gray shaded area.

## Discussion

Here we examined how genetic predisposition for ASD relates to thalamic functional connectivity in infants 37–44 weeks postmenstrual age and to behavioral outcomes at 18 months of age. Remarkably, we find that ASD PGS has early impacts on brain network functional connectivity soon after birth and is predictive of elevated ASD-associated symptomatology, slower motor development, and more ADHD-related problems in toddlerhood. Across male and female infants, we find that higher polygenic liability for ASD is associated with weaker thalamic connectivity to posterior parietal cortex. Sex interaction analyses revealed that thalamic connectivity with the parietal cortex, temporal cortex, and insula is differentially associated with ASD PGS in males and females. In sex-stratified analyses, females demonstrated positive associations between ASD polygenic scores (PGS) and thalamic connectivity with parieto-cingulate and anterior temporal cortices, as well as a negative association with the parieto-occipital cortex, whereas in males, we observed a positive association between ASD PGS and thalamic connectivity with sensorimotor cortex. Our findings indicate that genetic liability for ASD has sex-specific effects on functional brain networks during the neonatal period, a time marked by dramatic brain development, potentially laying the foundation for altered neurodevelopmental trajectories that may manifest in later behavioral challenges.

Our findings show that genetic predisposition for ASD adversely affects behavioral development in toddlers. While the earliest behavioral signs of ASD often manifest around one year of age [48], formal clinical diagnoses are typically not made until preschool age [2]. As prior work has shown that early intervention can dramatically improve behavioral outcomes in ASD [49], [50], there is a pressing need to identify tools to inform earlier detection of youth likely to acquire a diagnosis. Here, we show that genetic liability for ASD predicts greater ASD-related traits as assessed by the Q-CHAT and worse motor development on the Bayley-III at 18 months, suggesting that ASD common variants have discernible effects on behavior. Although the Q-CHAT only broadly assesses ASD symptoms at this early age, our findings are consistent with prior work showing that ASD PGS are associated with elevated ASD-related traits at 6 years of age [21]. The relationship between ASD PGS and motor development also corroborates previous work demonstrating that ASD PGS are linked to atypical motor development, including lower muscle tone between 9 and 20 weeks of age [20], as well as reduced motor development at 18 months of age [21]. These links between genetic liability and early motor differences highlight motor functioning as an early and sensitive marker of atypical neurodevelopment. Indeed, motor impairments are prevalent in ASD [51] and are detectable in infants at elevated familial likelihood for a diagnosis [52], [53], [54], [55]. Altogether, these findings suggest that genetic liability for ASD confers broad risk for ASD-related traits and may exert early effects on motor development.

Consistent with this pattern, we also find that higher ASD PGS is associated with greater ADHD problems at 18 months of age. ASD and ADHD are highly heritable and frequently co-occurring neurodevelopmental disorders, with approximately 40% of autistic individuals also acquiring an ADHD diagnosis [56]. A substantial body of evidence indicates that ASD and ADHD demonstrate considerable genetic pleiotropy, including contributions from both rare and common variants [16], [57], [58], [59], [60], [61], [62]. Our findings suggests that common genetic variation predisposing to ASD can help inform early detection of ADHD symptomatology in toddlerhood and underscores the potential shared genetic etiologies underlying these two neurodevelopmental conditions. This highlights that ASD-associated polygenic liability can shape early emerging behavioral trajectories beyond ASD-specific features, extending to broader domains of attention and regulation.

Alterations in thalamocortical functional connectivity have frequently been reported in ASD [13], [63], [64], [65], [66], [67]. The development of thalamocortical connectivity generally follows a sensorimotor-association axis [68], with the earliest connections forming between the thalamus and primary sensory processing areas and later-developing circuits supporting the complex transformation of sensory inputs to behaviors. A major hypothesis of the etiology of ASD is that perturbations in this developmental trajectory very early in life may alter the development of large-scale networks, setting the stage for atypical behavioral phenotypes observed across the autism spectrum [69]. Here, we show that across male and female infants, increased genetic liability for ASD is associated with weaker connectivity between the thalamus and posterior parietal cortex at 37-44 weeks postmenstrual age. As the posterior parietal association areas play an important role in combining and transforming sensorimotor inputs into coordinated cognitive and motor outputs [70], weaker thalamic connectivity with this region may reflect atypical development of higher-order integrative networks. Indeed, these findings are in line with prior reports of thalamic underconnectivity with the parietal cortex in children and adolescents with ASD [63], [71]. Altogether, our findings suggest that because genetic liability for ASD predicts differences in thalamocortical connectivity in infancy, previously observed underconnectivity with parietal regions in adolescent youth likely reflects an early neurodevelopmental pathway through which genetic liability for ASD manifests in the brain, rather than a consequence of ASD symptoms.

Sex differences in ASD diagnoses are well documented [72], yet it remains unclear how polygenic liability for the disorder may differentially impact the establishment of functional brain connectivity in male and female infants. Here, we find that sex interacts with ASD polygenic liability to shape thalamocortical organization, with distinct patterns emerging across sensory integration and salience-related regions. We observed strengthening of thalamo-parietal connectivity as a function of increased polygenic liability in males, but the opposite pattern of association in females. This suggests that while genetic risk for ASD amplifies early connectivity with cortical sensory regions in males, this increased genetic load may dampen connectivity of these same pathways in females. In contrast, the superior temporal gyrus and insula show the opposite pattern: higher ASD PGS predicted weaker thalamic connectivity in males and stronger connectivity in females, suggesting that in the face of elevated genetic risk for ASD, females may recruit higher-order circuits involved in language and salience detection [73] [74], while this connectivity pattern is weakened in males. Indeed, prior work has revealed sex differences in infant functional connectivity, with females showing stronger connectivity between temporal and frontal regions [75], as well as stronger connectivity between temporal language areas than males [76]. Taken together, our findings suggest that ASD genetic liability may interact with underlying sex differences in neurobiology to shape functional brain networks in neonates.

In sex-stratified analyses, we find sex-specific associations across primary sensory processing and sensory integration/association areas. Here, higher PGS was associated with stronger thalamic connectivity with the sensorimotor cortex in male infants. This finding may be linked to altered sensory processing often observed in ASD, typically manifesting in extreme aversions or reactions to sensory stimuli [77]. Prior research has shown that individuals with ASD display hyperconnectivity between the thalamus and precentral/postcentral gyri [65], [66] and show reduced modulation of this connectivity during sensory stimulation [12]. Consistent with these findings, we show that in male infants, greater genetic predisposition for ASD was associated with stronger thalamic connectivity to the sensorimotor cortex. This increased connectivity may reflect a decrease in thalamic gating to sensory processing regions of the cortex, ultimately resulting in increased excitation of primary sensorimotor cortices and leading to difficulties in the filtering and integrating of sensory information. Potential sex-differences in thalamic neurobiology have also been demonstrated by studies using magnetic resonance spectroscopy (MRS) to measure neurometabolites, which have found higher thalamic GABA concentrations in autistic males relative to females, indicating greater inhibitory signaling within the thalamus of male adults with ASD [78]. In this context, our finding of stronger thalamic connectivity to primary sensory processing regions in males with greater genetic liability for ASD may reflect underlying deficits in thalamic inhibitory mechanisms.

Finally, we report female-specific associations between genetic predisposition for ASD, thalamocortical connectivity, and later motor skills. Our findings show that higher PGS is associated with both weaker thalamic connectivity to the posterior parieto-occipital cortices and increased connectivity to parieto-cingulate and anterior temporal cortices in females. This corroborates prior reports of thalamic underconnectivity to parieto-occipital cortices and thalamic overconnectivity to temporal cortices in autistic children and youth [12], [13], [63], [64] and suggest that these patterns may be genetically driven and detectable early in life. Furthermore, in female infants, stronger thalamic connectivity with the anterior temporal cortex was associated with slower motor development, while stronger thalamic connectivity with the posterior parietal cortex was associated with more developed motor skills. This suggests that female patterns of connectivity linked to higher ASD genetic liability are associated with poorer motor development. This female-specific relationship between ASD-related connectivity and motor outcomes may reflect known sex differences in motor developmental trajectories in infancy and childhood, with females showing more advanced fine motor [79], [80] and gross motor skills [81] than males. Given this sex difference in the timing of motor development, our brain-behavior results may reflect functional connectivity patterns in females that are more tightly coupled to observable variation in motor behaviors in early toddlerhood, whereas these may not yet be linked to detectable motor differences in males. Overall, these findings suggest that polygenic liability for ASD shapes neonatal brain development in a sex-differentiated manner that has implications for later motor behaviors.

### Limitations

This study has several limitations. First, because the GWAS used to generate ASD PGS was restricted to European individuals, our analyses only included infants of European ancestry. Future progress in generating a multi-ancestry ASD GWAS will greatly expand the ancestry profile of individuals who can be included in subsequent analyses. Second, our functional connectivity analyses included only term-born infants due to known effects of preterm birth on brain development [82]. However, given the link between preterm birth and ASD diagnosis [83], additional studies on preterm-born infants may increase our understanding of how the interaction between ASD genetic predisposition and preterm birth may differentially impact brain functional connectivity and risk for ASD. Third, our seed-based fMRI analyses treated the thalamus as a unitary brain region due to a lack of individual thalamic nuclei parcellations in existing neonatal brain atlases. However, the thalamus consists of many specialized nuclei, each with specific cortical projections [64]. The development of neonate-specific thalamic atlas parcellations will allow for more granular investigations of thalamocortical functional connectivity. Finally, while this work shows that genetic predisposition for ASD impacts early thalamocortical functional connectivity within the first month of life, it remains unclear how this influences the longitudinal development of these brain functional networks. Future work leveraging larger, longitudinal datasets, such as the newly released HEALthy Brain and Child Development (HBCD) study [84] will be critical to fully understand the impact of ASD-associated variants on functional brain development.

### Conclusions

In summary, we show that genetic liability for ASD impacts neonatal thalamocortical connectivity and predicts suboptimal behavioral outcomes in toddlers. Our findings on the early emergence of sex-specific effects on thalamocortical connectivity add to the literature on sex differences in functional networks in ASD [26], [27], [28], [29] and importantly, highlight the role of ASD-related common genetic variation in the interplay between sex and functional brain connectivity during early life. While ASD PGS is an imperfect measure of ASD genetic liability, a deeper understanding of how genetic predisposition for ASD can shape brain and behavioral development is critical in informing more timely interventions.

## Data Availability

All data are available at National Institute of Mental Health (NIMH) Data Archive Collection #3955.

## Acknowledgements

We are grateful for the families and children who participated in the Developing Human Connectome Project (dHCP). The current project was supported by the National Institute of Mental Health at the National Institutes of Health grant number R01MH117982 to M.D., F31MH135704 to E.C., and K01MH135289 to L.M.H, and the Simons Foundation Autism Research Initiative grant number 00011789 to L.M.H.

