## Supplemental Table 1 for "Effects of polygenic liability for autism on neonatal thalamocortical connectivity and behavioral outcomes across sex"

Table S1. Coordinate table for functional connectivity peaks in ASD PGS regressions.

|  |  |  | **Peak (voxel coordinates)** | | | |
| --- | --- | --- | --- | --- | --- | --- |
|  | **Cluster coverage area** | **L/R** | **Max Z** | **x** | **y** | **z** |
| Full Sample PGS Association | **Parietal lobe** | R | 3.6 | 18 | -34 | 39.5 |
| Male > Female Interaction | **Parietal lobe** | R | 3.96 | 7.97 | -50 | 47.5 |
| Female > Male Interaction | **Superior temporal gyrus**, anterior temporal lobe, insula | L | 3.43 | -28 | -6.03 | 1.49 |
| Male PGS Positive Association | **Frontal lobe**, parietal lobe | L | 3.4 | -12 | -18 | 49.5 |
| Female PGS Positive Association | **Parietal lobe**, cingulate gyrus | R | 3.69 | 5.97 | -40 | 27.5 |
|  | **Anterior temporal lobe** | L | 4.14 | -16 | -4.03 | -12.5 |
| Female PGS Negative Association | **Parietal lobe** | R | 3.55 | 20 | -38 | 35.5 |
|  | **Occipital lobe** | R | 3.5 | 18 | -56 | 21.5 |

Regions listed in bold are peaks. Additional regions included in the cluster beyond the peak are also described.
