## Supplemental Table 2 for "Effects of polygenic liability for autism on neonatal thalamocortical connectivity and behavioral outcomes across sex"

Table S2. Summary of regression statistics for full sample and each sex separately.

|  | **Coefficient** | **SE** | **P-value** |
| --- | --- | --- | --- |
| Q-CHAT Total | 0.91 | 0.46 | 0.04* |
| Males | 0.97 | 0.78 | 0.21 |
| Females | 0.77 | 0.52 | 0.14 |
| Bayley-III Cognitive Composite | -0.31 | 0.57 | 0.59 |
| Males | -0.50 | 0.88 | 0.57 |
| Females | -0.08 | 0.75 | 0.92 |
| Bayley-III Motor Composite | -1.27 | 0.56 | 0.02* |
| Males | -1.61 | 0.90 | 0.08 |
| Females | -1.14 | 0.71 | 0.11 |
| Bayley-III Language Composite | -0.47 | 0.82 | 0.56 |
| Males | -1.05 | 1.31 | 0.42 |
| Females | 0.24 | 1.03 | 0.82 |
| CBCL ADHD T-Score | 0.01 | 0.01 | 0.03* |
| Males | 0.02 | 0.01 | 0.06 |
| Females | 0.01 | 0.01 | 0.22 |
| CBCL Anxiety Problems T-Score | 0.01 | 0.005 | 0.21 |
| Males | 0.01 | 0.01 | 0.19 |
| Females | 0.002 | 0.01 | 0.74 |
| CBCL Depressive Problems T-Score | 0.003 | 0.004 | 0.50 |
| Males | -0.001 | 0.005 | 0.81 |
| Females | 0.004 | 0.01 | 0.46 |
| CBCL Autism Spectrum Problems T-Score | 0.002 | 0.005 | 0.70 |
| Males | -0.002 | 0.007 | 0.75 |
| Females | -0.003 | 0.006 | 0.59 |
| CBCL Oppositional Defiant Problems T-Score | 0.007 | 0.005 | 0.20 |
| Males | 0.01 | 0.01 | 0.17 |
| Females | 0.01 | 0.01 | 0.46 |

Statistics (unstandardized coefficients, standard error (SE), and P-values) were derived from linear regression models assessing the relationship between ASD PGS and behavioral outcomes at 18 months in the full sample, and in males and females separately. Statistical significance is denoted with an asterisk (*). Abbreviations: QCHAT: Quantitative Checklist for Autism in Toddlers; Bayley-III: Bayley Scales of Infant and Toddler Development; CBCL: Child Behavior Checklist for Ages 1.5-5
